# Effect of a Dietary Intervention on Weight Loss in Adults with High vs Low Genetic Predisposition to Higher BMI: A Randomized Diet Intervention Trial

**DOI:** 10.1101/2025.10.06.25337395

**Authors:** Rodosthenis S. Rodosthenous, Leena E. Viiri, Anne Carson, Tatiana Cajuso, Andrea Corbetta, Samuel Jones, Ettore Brenna, FinnGen, Enni Makkonen, Minttu Virolainen, Niina Korkeala, Elisabeth Widén, Samuli Ripatti, Andrea Ganna

**Author notes:** Corresponding author: Name: Andrea Ganna, E-mail address, Postal address: Institute for Molecular Medicine Finland HiLIFE, University of Helsinki, Biomedicum 2U, Tukholmankatu 8, 00290 Helsinki.

## Abstract

It is unclear whether the effectiveness of weight loss interventions differs based on genetic predisposition to higher body mass index (BMI). Genome-wide polygenic scores (PS) for BMI are the strongest genetic predictors of elevated body weight, but no prospective trials have evaluated weight loss responses based on BMI PS. To assess whether adults with high or low genetic predisposition to higher BMI experience differential weight loss during a 6-month dietary intervention, we designed the GENEROOS study, a single-site, randomised controlled trial conducted in Finland from October 2023 to November 2024. Participants (N=223), aged 30–65 years, with a BMI of 23–36 kg/m² and no diabetes, were recruited from 38,621 genotyped individuals in the Finnish Clinical Biobank Tampere. They were selected from the top or bottom 5th percentile of the BMI PS distribution and randomized to either a six-month dietary coaching program aimed at a 500 kcal/day energy deficit or to a control group receiving no dietary intervention. The main outcome was change in body weight (%) from baseline to 6 months and the interaction between BMI PS and intervention status. At baseline, those in the top 5% of BMI PS weighed on average 8.4 kg (95% CI, 5.1 to 11.7 kg) more than those in the bottom 5%. In the intervention group, mean weight change at 2, 4, and 6 months was −3.89%, −5.32%, and −4.70%; in the control group it was −1.10%, −0.85%, and 0.14%, respectively. No significant interaction was found between BMI PS and intervention on weight change (β = 0.06 [95% CI, −1.33 to 1.44]; P = .94). In this randomised trial, genetic predisposition as measured by BMI PS did not modify response to dietary intervention, indicating that leveraging polygenic scores may not enhance the effectiveness of personalized weight loss strategies.

**Trial Registration:** ClinicalTrials.gov Identifier: NCT06092372

## INTRODUCTION

The prevalence of overweight and obesity is increasing globally, and it is a major public health concern. By 2035, 54% of the global adult population is estimated to be either overweight (body mass index (BMI) ≥ 25 kg/m2) or obese (BMI ≥ 30 kg/m^2^)^1^. High BMI is a major risk factor for several chronic diseases including cardiovascular disease^2^, diabetes mellitus, chronic kidney disease^3^ and certain cancers^4^ which can lead to premature mortality.

Overweight and obesity are complex, multifactorial conditions influenced by both genetic and environmental factors. Environmental factors contributing to a high BMI include unhealthy diet, low physical activity, and sedentary behaviors^5,6^. Despite environmental factors playing an important role, the heritability of BMI is estimated to be moderate to high, ranging from 0.23 to 0.90 depending on study design, age group, and population^7–9^. Twin and family studies consistently demonstrate substantial genetic contribution to BMI variability, with higher heritability estimates in children compared to adults, suggesting that genetic factors may exert a stronger influence early in life^7^. While rare monogenic forms of obesity exist, most genetic susceptibility arises from the cumulative effects of common single nucleotide polymorphisms (SNPs). To date, over a thousand SNPs have been associated with BMI^10–12^. Polygenic scores (PS), which aggregate the effects of thousands of SNPs, explain approximately 8.4% of BMI variation^8^, capturing only a fraction of the heritability estimated from twin and family studies^7^. In the UK Biobank, individuals in the top PS decile had a 25-fold higher risk of severe obesity (BMI > 40 kg/m²) compared to those in the bottom decile^8^. However, PS is not deterministic, emphasizing that while genetic susceptibility plays a significant role, the interplay with the environment is crucial in shaping BMI over a lifetime.

Numerous dietary intervention studies have aimed to reduce body weight in overweight and obese individuals. A meta-analysis of clinical trials reported a mean weight loss of 5 to 8.5 kg (5% to 9%) during the first 6 months following reduced-energy diet interventions^13^. While the role of genetic variation in diet response remains unclear, some studies have suggested that genetic factors may influence individual responses to dietary and lifestyle interventions, while others not^14^. For example, a meta-analysis including retrospective analysis of diet interventions, found that individuals carrying the homozygous FTO obesity-predisposing allele experienced greater weight loss through diet and lifestyle changes compared to non-carriers^15^. On the contrary, a meta-analysis by Livingstone et al., found that carriers of the FTO minor allele had a similar response to diet, drug or lifestyle interventions compared to non-carriers^16^. To the best of our knowledge, the DIETFITS study is the only randomized trial that prospectively tested whether a three-SNP genetic score modified weight change in response to a healthy low-fat versus a healthy low-carbohydrate diet, but no significant differences were observed^17^.

Genome-wide PS offer a substantially more powerful measure of genetic risk for BMI than individual SNPs such as those in the FTO gene. Two large behavioral intervention trials have investigated BMI polygenic scores or obesity-associated SNPs in relation to weight loss outcomes. Papandonatos et al. evaluated 91 BMI-associated SNPs from the Diabetes Prevention Program (DPP) and Look AHEAD trials, finding no consistent evidence that genetic risk predicted weight loss or regain^18^. More recently, the ONTIME study by Dashti et al. employed a genome-wide polygenic score and identified that higher BMI PS was associated with a slower rate of weight loss during a structured behavioral intervention, though no association with total weight loss or interaction with intervention intensity was found^14^. However, existing studies have been limited by observational designs, absence of control groups or lack of targeted recruitment based on genetic risk.

The GENEROOS study was designed to fill this gap by prospectively testing whether BMI polygenic risk affects weight loss following a dietary intervention. To further maximize power, we used a biobank-based genetic-first recontact strategy, recruiting participants at the extremes of BMI genetic risk from a pool of 38,621 already-genotyped individuals as part of FinnGen study^19^. The primary objective of the GENEROOS study was to test whether overweight or mildly obese non-diabetic individuals in the top and bottom 5^th^ percentile of BMI PS show differential success in 6-month weight change due to dietary and lifestyle intervention, compared to controls.

## RESULTS

Among 38,621 genotyped participants, 959 met initial eligibility criteria and were invited to the study, 529 in the Top 5% and 430 in the Bottom 5% of the BMI PS. Out of those invited, 642 (66.9%) did not respond, 79 (8.2%) responded but did not meet the study eligibility criteria, and 15 (1.6%) did not enroll in the study, resulting in 223 participants (23.3%) completing the first in-person visit and being randomized (mean age, 51.1 [SD, 10.4] years; 74.9% women; mean body mass index, 30.3 [SD, 3.2]). The study enrolment rate was very similar to the 23.1% we observed in a recent biobank-based recall study in Finland^20^. The flow of the study population through the study is shown in **Figure 2**.

**Figure 1:**
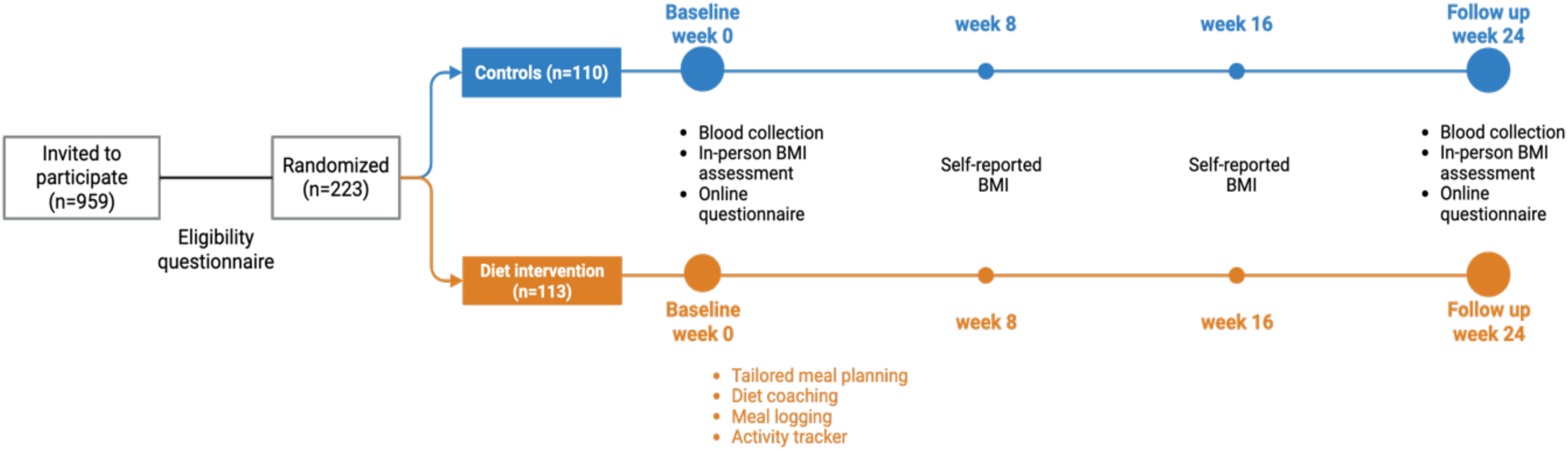
GENEROOS study design.

**Figure 2:**
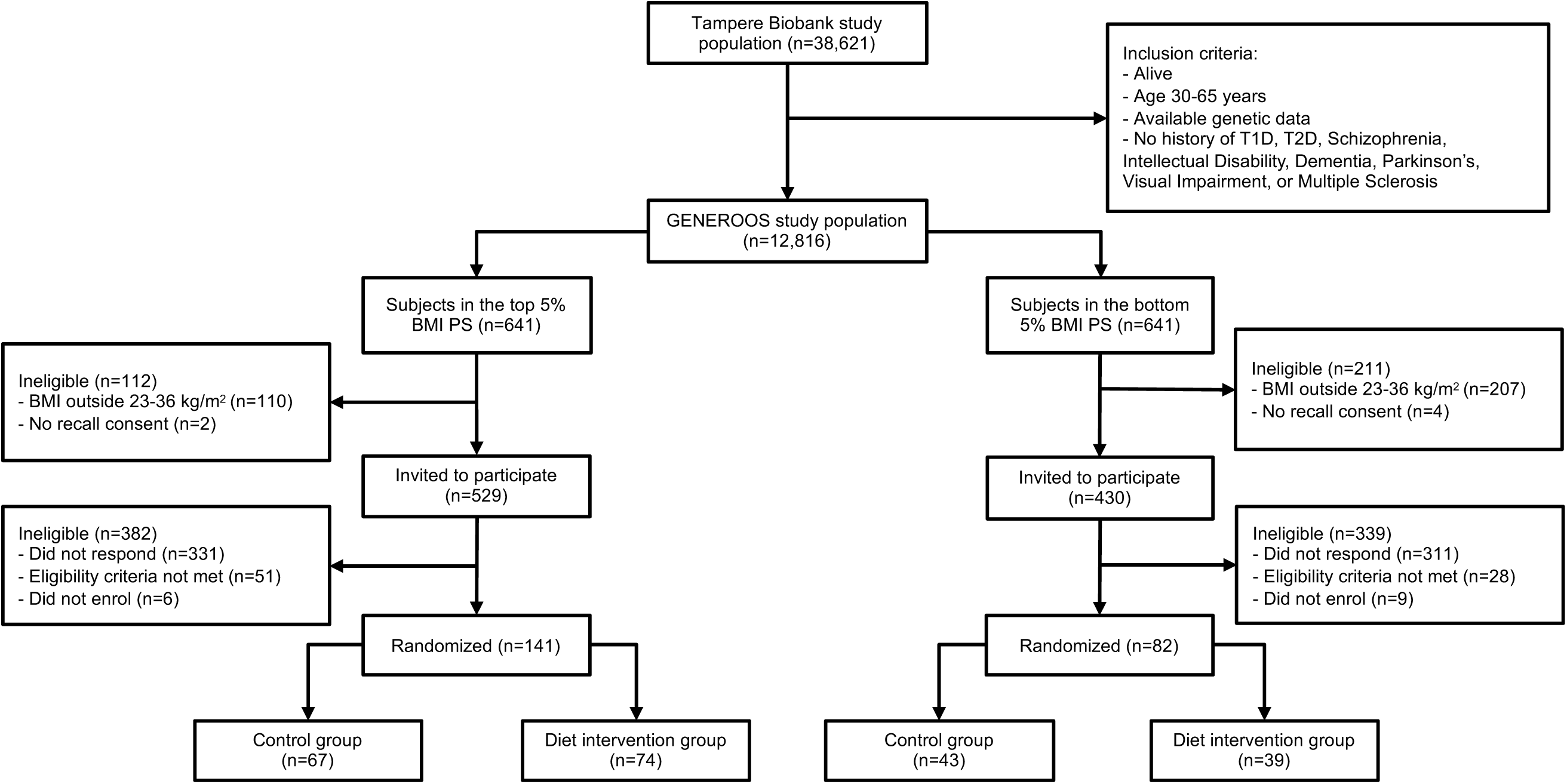
Flow diagram for GENEROOS study participants.

There was no difference in demographic, anthropometric and biomarkers characteristics between study arms at baseline (**Table 1**). While we observe overall more individuals belonging to the top 5% of BMI PS than in the bottom 5% (N=141 [63.2%] vs N=82 (36.8%)), the distribution of two groups was similar between study arms (**Figure 2**). At baseline, individuals in the top 5% of BMI PS had on average, 3.0 kg/m^2^ (95% CI, 2.2 to 3.8 kg/m^2^) higher BMI and 8.4 kg (95% CI, 5.1 to 11.7 kg)) higher body weight than those in the bottom 5% of BMI PS (**Suppl. Figure 2**).

**Table 1:**
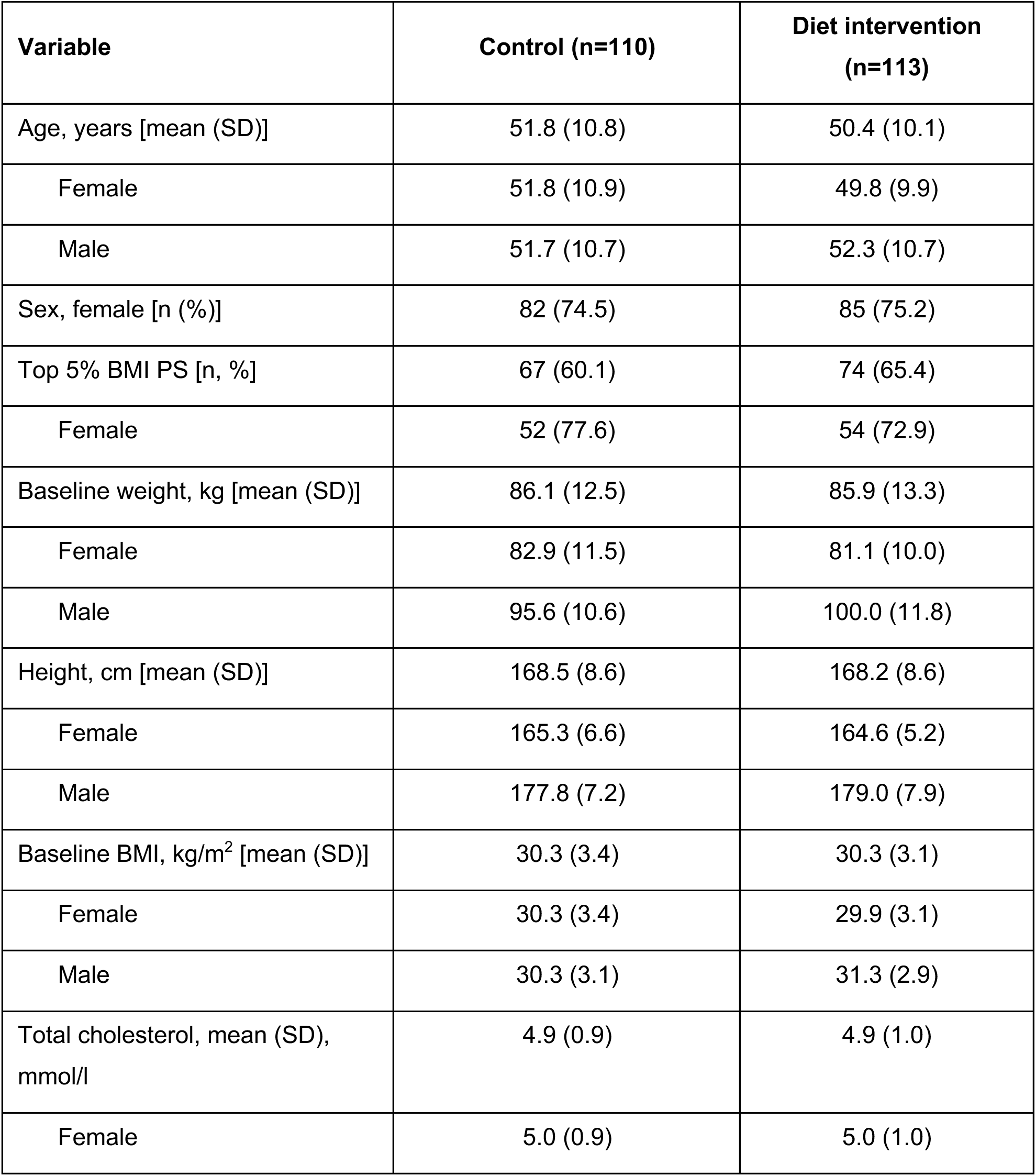
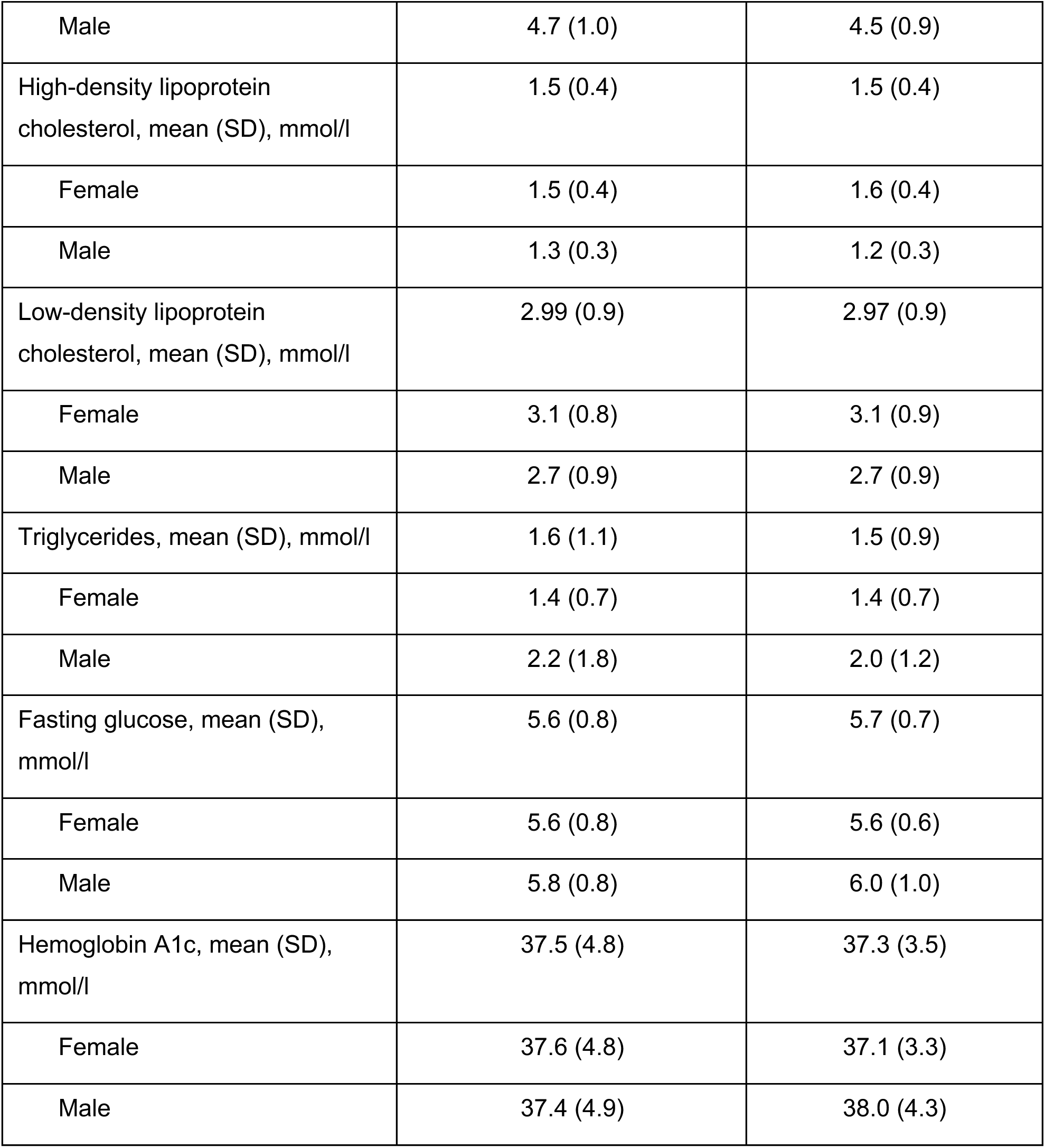
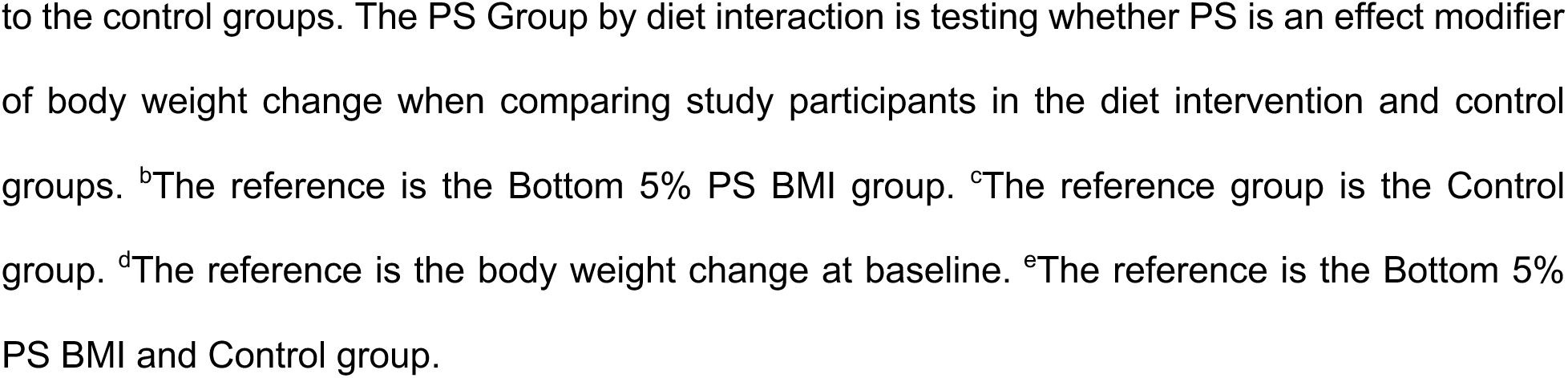
Baseline descriptive characteristics of the GENEROOS study participants.

### Primary Outcome

Of all individuals that participated in the baseline visit, 13 (5.8%) did not return to the 6 months follow-up visit. The proportion of lost at follow-up was similar between the diet intervention and control group (6.2% vs 5.5%). In the diet intervention arm, the mean change (%) in body weight at 2 months was −3.89 (95% CI, −4.40 to −1.97, n=109), at 4 months was −5.32 (95% CI, −6.18 to −4.46, n=108), and at 6 months (end of study) was −4.70 (95% CI, −5.78 to −3.61, n=106), whereas in the control group the mean change (%) in body weight at 2 months was −1.10 (95% CI, −1.49 to −0.70, n=107), at 4 months was −0.85 (95% CI, -1.41 to −0.29, n=106), and at 6 months (end of study) was 0.14 (95% CI, −0.54 to 0.82, n=104) (**Figure 3**). At 6 months, mean weight change (kg) was −4.02 kg (95% CI, −5.96 kg to −3.08 kg) for the diet intervention and 0.04 kg (95% CI, −0.57 kg to 0.65 kg) for the control group, which was statistically significantly different (p<0.001). On the contrary, the mean 6-month weight change (kg) by BMI PS group was not significantly different with −1.9 kg (95% CI, −3.1 kg to −1.3 kg) in the top 5% BMI PS and −2.2 kg (95% CI, −2.8 kg to −1.1 kg) in the bottom 5% BMI PS. Further, we did not observe a statistically significant interaction (P=0.94) between the BMI PS group and diet intervention when using linear mixed models including self-reported weight measurements at month 2 and 4 (**Table 2**).

**Figure 3:**
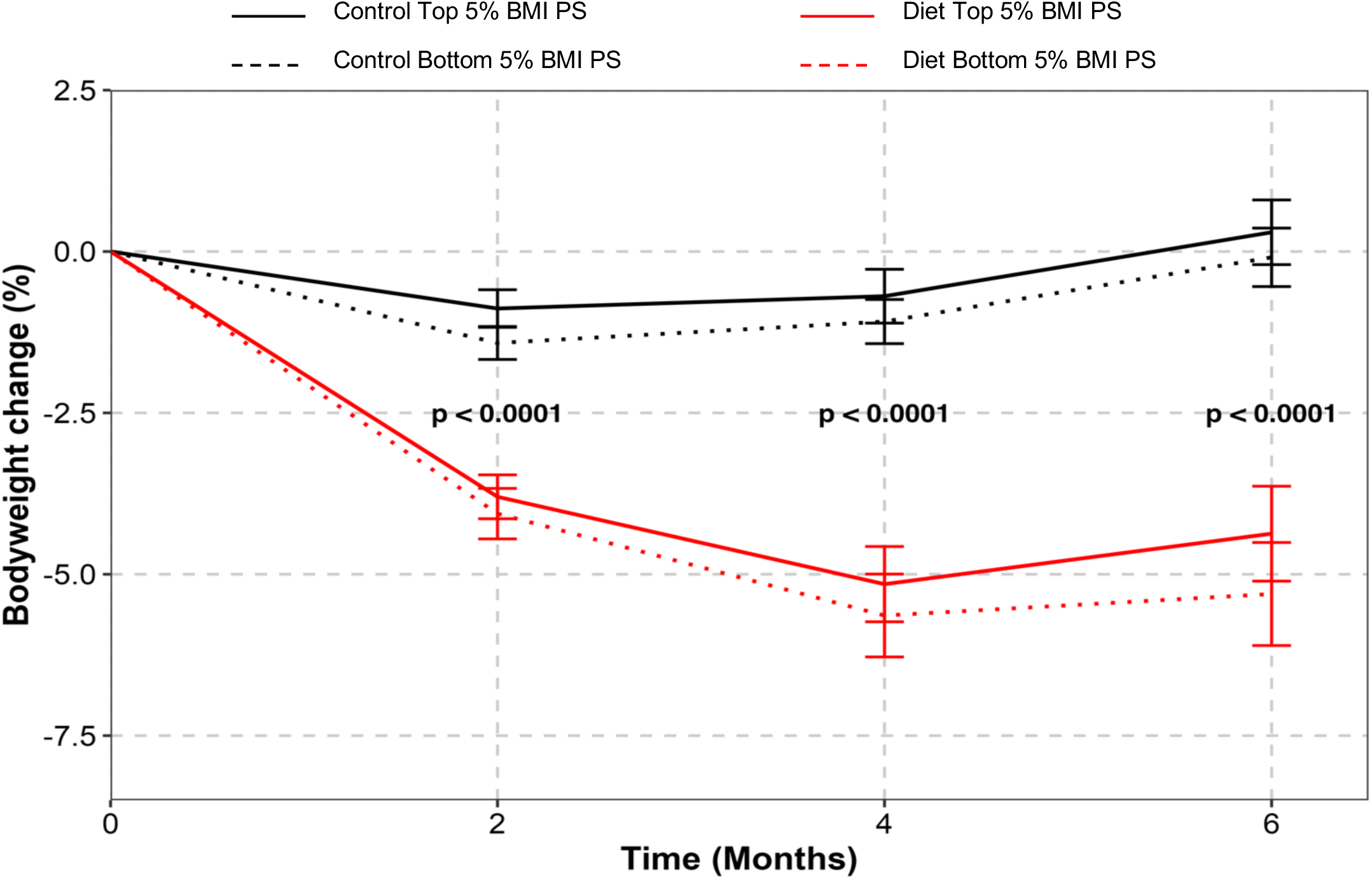
Change in body weight (%) between the control and diet intervention by the PS group (N=223). P-value at each time point was calculated using Mann-Whitney U (Wilcoxon rank-sum) non-parametric test.

**Table 2:**
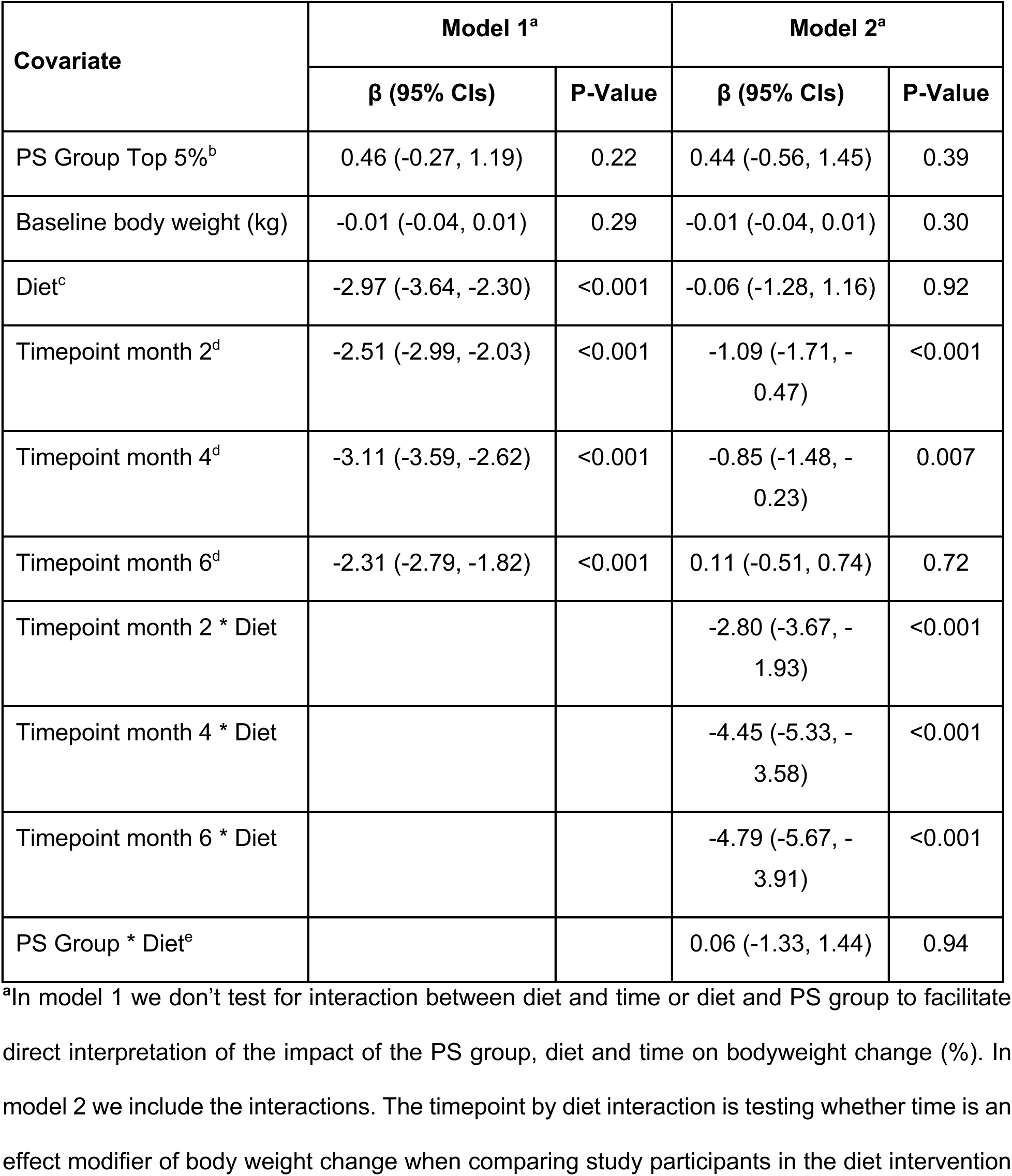
Association between bodyweight change (%) and BMI PS among study participants from linear mixed effects models (N_individuals_ = 223 and N_observations_ = 863). All covariates used in the model are reported.

### Non-prespecified exploratory analysis

Individuals in the diet intervention group compared to controls showed lower levels (mean, 95% CI, mmol/l) of total cholesterol (−0.205, 95% CI: -0.329 to -0.081 vs -0.009, 95% CI: -0.141 to 0.122), LDL cholesterol (−0.237, 95% CI: -0.351 to -0.122 vs -0.056, 95% CI: -0.171 to 0.059), triglycerides (−0.218, 95% CI: -0.360 to -0.075 vs -0.080, 95% CI: -0.236 to 0.076), HbA1C (−0.623, 95% CI: -0.988 to -0.257 vs -0.058, 95% CI: -0.691 to 0.575), glucose (−0.142, 95% CI: - 0.257 to -0.026 vs -0.003, 95% CI: -0.113 to 0.107), but not HDL cholesterol (−0.018, 95% CI: - 0.053 to 0.017 vs -0.008, 95% CI: -0.042 to 0.026) (**Suppl. Fig. 3**).

## DISCUSSION

It remains unclear whether genetic factors contribute to individual differences in weight loss following dietary interventions. In this prospective randomized study of overweight and mildly obese, non-diabetic individuals with extreme high or low genetic risk for BMI, a six-month dietary intervention effectively reduced body weight (−4.7%) and improved key cardiometabolic biomarkers, including LDL cholesterol, triglycerides, glucose, and HbA1c, regardless of genetic risk group. However, weight loss was similar across genetic risk groups, suggesting that genetic predisposition to high BMI, as measured by a genome-wide polygenic score does not significantly modify the effectiveness of diet and lifestyle interventions for weight reduction.

Two previous meta-analyses^15,16^ have reported conflicting results on the differential impact that diet interventions have on FTO risk allele carriers. BMI PS capture a substantially larger proportion of genetic risk for BMI compared to single variants such as FTO – which is also included in the PS - providing greater power to detect differential dietary effects. In our study, the differences in BMI between high and low PS groups (31.3 kg/m^2^ vs. 28.4 kg/m^2^) were substantially larger than what reported between carriers and non-carriers of the homozygous FTO risk allele (an increase of 0.31 kg/m^2^ (0.14 to 0.47); P<0.001) for each additional copy)^16^. It remains possible, however, that the biological pathways linked to specific variants like FTO may influence diet responsiveness differently from the broader genetic effects captured by BMI PS.

Our findings are concordant with prior large-scale studies suggesting limited or no influence of genome-wide BMI polygenic scores on weight loss response to behavioral interventions. In the Look AHEAD and Diabetes Prevention Program (DPP) trials, only one of 91 known BMI-associated variants (MTIF3-rs1885988) showed an association with total weight loss, underscoring a minor role of individual genetic variants in modifying intervention outcomes. Similarly, in the ONTIME study—a genome-wide analysis of behavioral weight loss in over 1,200 individuals—a higher BMI polygenic score was associated with a slightly attenuated rate of weight loss, but not with total weight loss or final outcomes. It is noteworthy that the weight loss achieved through the dietary intervention (−4.02 kg) was approximately half of the baseline difference in body weight between BMI PS extremes (8.23 kg). This suggests that, although the intervention was equally effective across genetic risk groups, it was not sufficient to overcome the underlying genetic differences in body weight between the PS extremes.

Our study has several limitations. First, there was a higher proportion of women (74.9%) than men, likely due to known sex differences in volunteer participation, though diet effectiveness was similar across sexes. Second, only 36.8% of participants were from the bottom 5th percentile of BMI PS, as many were excluded for having BMI below 23 kg/m² at screening. Third, we did not collect detailed calorie intake data, limiting insights into behavioral mechanisms of weight loss; however, improvements in cardiometabolic biomarkers suggest meaningful physiological changes beyond weight loss. Fourth, although we used the most advanced BMI PS available at the time, it does not fully capture genetic contributions to BMI. Future studies should test whether newer, more predictive scores replicate these findings. Finally, generalizability may be limited to populations with the resources, interest, and digital literacy to engage in a structured dietary intervention.

A major strength of this study is its prospective, randomized design, in contrast to retrospective analyses often used to evaluate genetics in dietary response. Using a genetic-first recontact strategy from a cohort of over 38,000 genotyped individuals, we recruited participants at the extremes of BMI genetic risk, maximizing power to detect differential effects. Although our hypothesis was not supported, this approach shows how biobank-based genetic recruitment can accelerate precision medicine trials. Another strength is the inclusion of a non-intervention control group, the gold standard for testing dietary intervention efficacy. Despite the potential for higher dropout in control groups, follow-up loss was <5% and similar across groups, supporting the robustness of our findings. Finally, whereas most weight loss trials target individuals with obesity and co-morbidities (e.g., type 2 diabetes), we focused on overweight participants without diabetes (BMI 23–36), a population more representative of those who might benefit from preventive interventions.

In conclusion, this 6-month randomized dietary intervention study found no evidence that BMI polygenic scores predict differential weight loss outcomes among overweight and mildly obese non-diabetic individuals. Our findings suggest that, in this population, current genome-wide BMI polygenic scores are not helpful in identifying who would benefit most from dietary and lifestyle interventions aimed at reducing body weight and improving cardiometabolic health. These findings underscore the importance of critically evaluating genetic tools before integrating them into clinical decision-making.

## Data Availability

De-identified individual participant data underlying the results reported in this article, including data dictionaries, will be made available via the European Genome-Phenome Archive (EGA; https://ega-archive.org) by searching for the study ID (to be provided), requesting access, and getting approval by the GENEROOS study research team. The shared dataset includes all primary and secondary outcomes, baseline characteristics, and other variables used in the analysis. Data will be available beginning at the time of publication and for a period of five years. Access requests must include a methodologically sound proposal, will be subject to review within four weeks, and require a valid EGA account. Data reuse for commercial purposes or inclusion of external collaborators not named in the original proposal requires Steering Committee approval. All data will be de-identified in accordance with relevant regulations. Secondary users must agree not to attempt re-identification of participants and must cite the original trial (ClinicalTrials.gov: NCT06092372) and specify how their analyses differ from previously reported findings.

## ACKNOWLEDGMENTS

This study has received funding from the European Research Council (ERC) under the European Union’s Horizon 2020 research and innovation program (grant number 945733). We also want to acknowledge the participants and investigators of the FinnGen study. The FinnGen project is funded by two grants from Business Finland (HUS 4685/31/2016 and UH 4386/31/2016) and the following industry partners: AbbVie Inc., AstraZeneca UK Ltd, Biogen MA Inc., Bristol Myers Squibb (and Celgene Corporation & Celgene International II Sàrl), Genentech Inc., Merck Sharp & Dohme LCC, Pfizer Inc., GlaxoSmithKline Intellectual Property Development Ltd., Sanofi US Services Inc., Maze Therapeutics Inc., Janssen Biotech Inc, Novartis AG, and Boehringer Ingelheim International GmbH. Following biobanks are acknowledged for delivering biobank samples to FinnGen: Auria Biobank (www.auria.fi/biopankki), THL Biobank (www.thl.fi/biobank), Helsinki Biobank (www.helsinginbiopankki.fi), Biobank Borealis of Northern Finland (https://www.ppshp.fi/Tutkimus-ja-opetus/Biopankki/Pages/Biobank-Borealis-briefly-in-English.aspx), Finnish Clinical Biobank Tampere (https://www.pirha.fi/en/web/english/for-professionals/finnish-clinical-biobank-tampere), Biobank of Eastern Finland (www.ita-suomenbiopankki.fi/en), Central Finland Biobank (www.ksshp.fi/fi-FI/Potilaalle/Biopankki), Finnish Red Cross Blood Service Biobank (www.veripalvelu.fi/verenluovutus/biopankkitoiminta), Terveystalo Biobank (www.terveystalo.com/fi/Yritystietoa/Terveystalo-Biopankki/Biopankki/) and Arctic Biobank (https://www.oulu.fi/en/university/faculties-and-units/faculty-medicine/northern-finland-birth-cohorts-and-arctic-biobank). All Finnish Biobanks are members of BBMRI.fi infrastructure (https://www.bbmri-eric.eu/national-nodes/finland/). Finnish Biobank Cooperative - FINBB (https://finbb.fi/) is the coordinator of BBMRI-ERIC operations in Finland. The Finnish biobank data can be accessed through the Fingenious^®^ services (https://site.fingenious.fi/en/) managed by FINBB.

## AUTHOR CONTRIBUTIONS

Drs Rodosthenous and Ganna had full access to all of the data in the study and take responsibility for the integrity of the data and the accuracy of the data analysis.

**Concept and design:** Rodosthenous, Widén, Ripatti, Ganna

**Acquisition, analysis, or interpretation of data:** Rodosthenous, Viiri, Corbetta, Jones, Brenna, Makkonen, Virolainen, Korkeala

**Administrative, technical, or material support:** Carson, Cajuso, Viiri, Makkonen, Virolainen, Korkeala

**Statistical analysis:** Rodosthenous, Brenna

**Drafting of the manuscript:** Rodosthenous, Cajuso, Brenna, Ganna

**Critical review of the manuscript for important intellectual content**: All authors.

**Obtained funding:** Ganna

**Supervision:** Widén, Ripatti, Ganna

**Final approval of the version to be published:** All authors.

## METHODS

### Study design

The Role of Genetics in Weight Loss Study (GENEROOS, ClinicalTrials.gov: NCT06092372) was a 6-month, single-site, parallel-arm diet intervention trial (Figure 1, created in BioRender. Rodosthenous, R. (2025) https://BioRender.com/y4ujzed). The approved protocol, including the study design and statistical analysis plan can be found in Supplementary Material File 1. In brief, participants were enrolled between October 2, 2023, and November 8, 2024. Under the Finnish Biobank Act (688/2012), biobanks may recontact individuals who have consented to such contact. Eligible individuals for recontact had to be alive, have consented to recontact via the Finnish Clinical Biobank Tampere, have genetic data from FinnGen release 11, be in the top or bottom 5th percentile of the BMI PS distribution, aged 30–65 years, and have a BMI of 23–36 based on their latest electronic health record (EHR) from Tampere University Hospital. Those without a recorded BMI were also eligible. Using nationwide inpatient and outpatient registers, we excluded individuals with a diagnosis of Type I or II Diabetes (ICD-10: E10–E14), prior bariatric surgery (Operation codes: JDF00–01, JDF10–11, JDF20–21, JDF96–97, JFD03–04, JFD20, JFD96), schizophrenia or intellectual disability (ICD-10: F20–F29, F70–F79), dementia or Parkinson’s (ICD-10: F00–F09, G20, G30), visual impairment (ICD-10: H54), or multiple sclerosis (ICD-10: G35).

### Ethics statement

The GENEROOS study was approved by the Coordinating Ethics Committee of the Hospital District of Helsinki and Uusimaa (HUS/583/2022). No adverse effects, risks, or significant discomfort were anticipated for participants, aside from minor discomfort from blood draws. In accordance with Ethics Committee requirements, participants were informed about procedures for addressing any significantly abnormal laboratory findings that might require medical attention. Study subjects were also included in FinnGen and provided informed consent for biobank research, based on the Finnish Biobank Act. Details on the FinnGen study can be found in Supplementary Material File 2.

### Re-contacting approach

Eligible individuals were invited via physical letter from the Finnish Clinical Biobank Tampere. Interested participants logged into the study portal (https://omabiopankki.fingenious.fi/login), provided electronic informed consent, and completed an eligibility survey in RedCap^21^, which collected self-reported height, weight (for BMI calculation), pregnancy status, motivation to lose weight, and smartphone ownership (see Supplementary Material File 3) Those with a BMI between 23–36, a smartphone, interest in weight loss, and—if female—not pregnant or breastfeeding, were eligible to enroll and book a clinical lab visit.

All eligible participants were randomized to the control or diet intervention group using computerized randomization (randomizeR package, R version 2024.12.0+467). Group assignment was revealed by the research nurse only after the baseline lab visit. Participants were blinded to their PS group, and outcome assessors were blinded to PS group and lab measures, though not to intervention assignment.

### Blood sample collection and biomarker analysis

Fasting whole blood samples (3 x 3 mL) were collected by a trained bioanalyst in EDTA tubes (BD Vacutainer, catalog number 368499), heparin tubes (BD Vacutainer, catalog number 368497), and Vacuette glucose tubes (Greiner Bio-One, catalog number 454514), both at baseline and follow-up visits and shipped to Fimlab (Tampere, Finland) for biomarker analyses (i.e., total cholesterol, LDL, HDL, triglycerides, glucose and HbA1c). A non-prespecified exploratory analysis compared changes in all measured biomarkers between the diet and control groups. The results of biomarker analyses from both baseline and follow-up were returned to all study participants who provided informed consent and agreed to receive them at the end of the study. All biomarkers were analyzed in

### Assessment of the primary outcome

We calculated the participants’ BMI four times during the study: at baseline, two months and four months from enrollment and at the end of the study (Figure 1). At baseline and end of the study, participants’ BMI was calculated using the body weight and height as measured by a trained nurse in the clinic, whereas at months two and four was calculated using self-reported body weight and height via a text message. Participants were advised to weigh themselves in the morning, before breakfast, and after emptying their bladder. To ensure validity of self-reported BMI data at month two and four, we compared the baseline self-reported BMI collected from the online eligibility survey and in-person BMI measurements. The correlation between the two BMI measurements was high (Spearman’s rho = 0.93, **Suppl. Fig 1**).

### Polygenic score for body mass index

We derived a polygenic score (PS) for BMI using summary statistics from two sources: (1) Yengo et al., which performed a GWAS of BMI in approximately 700,000 genotyped individuals^12^, and (2) a GWAS of BMI in 315,152 individuals from FinnGen release 11, excluding participants from the Tampere biobank. The summary statistics from these two studies were meta-analyzed using METAL^22^. We then applied PRS-CS^23^, following the pipeline available at https://github.com/FINNGEN/CS-PRS-pipeline, to generate the PS weights from the meta-analyzed summary statistics. The PS was based on 1,097,993 SNPs. The polygenic scores were calculated for individuals in the Finnish Clinical Biobank Tampere using the -- score function in PLINK v2.00a4LM^24^. Among the 25,941 genotyped individuals with available BMI from the Finnish Clinical Biobank Tampere, the BMI PS explained 12.7% of the variance in BMI as measured by the coefficient of determination (R^2^).

### Diet intervention

The diet intervention coaching program, provided by ViaEsca Oy (Helsinki, Finland, viaesca.com), supported participants in reducing body weight and adopting a sustainable, balanced approach to healthy eating. It emphasized calorie intake and expenditure, using guidance based on official dietary recommendations. Interventionists were blinded to participants’ PS group and lab measures. A standardized approach ensured all participants began from the same baseline, based on the principle that behavioral change is required for outcomes like body weight reduction. The foundational program, regardless of prior habits, addressed eating rhythm, portion control, balanced meals, caloric needs tied to activity levels, and preparation of nutritious, simple meals. Participants received structured meal plans of five daily meals, personalized to individual caloric needs and set to 500 kcal/day below estimated energy expenditure. Plans followed macronutrient targets of 45–60% carbohydrates, 25–35% fat (at least two-thirds unsaturated), and 15–25% protein, including at least 35g fiber and 500– 800g of vegetables and fruits daily.

Caloric needs were estimated from registration data (age, height, weight, gender, and activity level), and refined with linked activity trackers using a rolling 7-day calorie average whenever possible. Participants could follow or adapt the plan to meet their energy target, by voluntarily logging meals in the ViaEsca mobile app to monitor intake. A food diary allowed adjustments for meals outside the plan. Two automated messaging types supported participants: time-based messages with daily tips, educational content, and progress check-ins, and activity-based messages providing real-time feedback on intake and prompting coach interaction if targets weren’t met. Participants also had online coaching access via chat for support and guidance, if needed.

### Statistical analysis

Based on the original study design with 62 participants in each of the 4 relevant groups (Top/Bottom 5% BMI PS and dietary intervention), and assuming normally distributed weight change at 6 months, there was 80% power to detect a 15% difference in diet intervention effect by BMI PS group (i.e., whether diet intervention affected weight change between BMI PS groups). This was based on simulations and a 2-sided Wald test at the .05 significance level.

The primary outcome was body weight change (%) at 6 months; all other measures were secondary. Outcomes are presented as mean and 95% confidence intervals (CI). An intent-to-treat principle was applied, including all randomized participants with baseline data in the analysis according to original assignment, regardless of adherence or follow-up. This assumes missing follow-up data are unrelated to unobserved weight values, conditional on treatment assignment and baseline/intermittent weights.

Linear mixed-effects models for repeated measures were used to assess the effects of diet intervention, PS group, and their interaction, using the *lme* function from the *nlme* package in R. Participants had random intercepts to account for individual-level variability. The primary test of interest was the interaction effect. To model within-subject correlation over time, unstructured, compound symmetry (CS), first-order autoregressive, and Toeplitz structures were evaluated.

The final structure (CS) was selected based on convergence and highest Bayesian Information Criterion (BIC). Parameters were estimated via restricted maximum likelihood (REML), and missing data were handled under the missing at random (MAR) assumption using na.exclude. Standard diagnostics assessed normality and homoscedasticity of residuals. Differences in body weight and BMI, between Top 5% vs Bottom 5% BMI PS at baseline were compared using the Mann-Whitney U (Wilcoxon rank-sum) test.

Non-prespecified exploratory analyses comparing changes in total cholesterol, LDL, HDL, triglycerides, glucose, and HbA1c between diet and control groups were conducted using the Mann-Whitney U (Wilcoxon rank-sum) test. All analyses were conducted in RStudio (version 2024.12.0+467).

### Changes compared to the registered trial protocol

Compared to the submitted trial protocol, there have been two major changes. First, we originally planned to recruit 300 participants per PS group across three biobanks. Due to logistical challenges, we instead focused on a single biobank and recruited fewer participants than originally planned. The updated power calculations reflect this revised scenario. Second, while the original protocol stated that we would measure clinical biomarkers, we did not explicitly specify them as a “secondary outcome.” Because of this, we refer to these analyses in the manuscript as a “non-prespecified exploratory analysis.”

## CODE AVAILABILITY STATEMENT

R code used for data preparation and analysis can be found at the GENEROOS GitHub repository: https://github.com/dsgelab/GENEROOS_Public.

## REFERENCES

1. Observatory WOFGO. World Obesity Federation Global Obesity Observatory World Obesity Federation Global Obesity Observatory Web site. Available at: https://data.worldobesity.org (Accessed: 18 February 2025). Published 2025. Accessed.

2. Emerging Risk Factors C, Danesh J, Erqou S, et al. The Emerging Risk Factors Collaboration: analysis of individual data on lipid, inflammatory and other markers in over 1.1 million participants in 104 prospective studies of cardiovascular diseases. Eur J Epidemiol. 2007;22(12):839–869.

3. Singh GM, Danaei G, Farzadfar F, et al. The age-specific quantitative effects of metabolic risk factors on cardiovascular diseases and diabetes: a pooled analysis. PLoS One. 2013;8(7):e65174.

4. Lauby-Secretan B, Scoccianti C, Loomis D, et al. Body Fatness and Cancer--Viewpoint of the IARC Working Group. N Engl J Med. 2016;375(8):794–798.

5. Nie Y, Zhang Y, Liu B, Meng H. Glucagon-Like Peptide-1 Receptor Agonists for the Treatment of Suboptimal Initial Clinical Response and Weight Gain Recurrence After Bariatric Surgery: a Systematic Review and Meta-analysis. Obes Surg. 2025;35(3):808–822.

6. Wells JC. Obesity as malnutrition: the role of capitalism in the obesity global epidemic. Am J Hum Biol. 2012;24(3):261–276.

7. Elks CE, den Hoed M, Zhao JH, et al. Variability in the heritability of body mass index: a systematic review and meta-regression. Front Endocrinol (Lausanne). 2012;3:29.

8. Khera AV, Chaffin M, Wade KH, et al. Polygenic Prediction of Weight and Obesity Trajectories from Birth to Adulthood. Cell. 2019;177(3):587–596 e589.

9. Maes HH, Neale MC, Eaves LJ. Genetic and environmental factors in relative body weight and human adiposity. Behav Genet. 1997;27(4):325–351.

10. Locke AE, Kahali B, Berndt SI, et al. Genetic studies of body mass index yield new insights for obesity biology. Nature. 2015;518(7538):197–206.

11. Loos RJF, Yeo GSH. The genetics of obesity: from discovery to biology. Nat Rev Genet. 2022;23(2):120–133.

12. Yengo L, Sidorenko J, Kemper KE, et al. Meta-analysis of genome-wide association studies for height and body mass index in approximately 700000 individuals of European ancestry. Hum Mol Genet. 2018;27(20):3641–3649.

13. Franz MJ, VanWormer JJ, Crain AL, et al. Weight-loss outcomes: a systematic review and meta-analysis of weight-loss clinical trials with a minimum 1-year follow-up. J Am Diet Assoc. 2007;107(10):1755–1767.

14. Dashti HS, Scheer F, Saxena R, Garaulet M. Impact of polygenic score for BMI on weight loss effectiveness and genome-wide association analysis. Int J Obes (Lond). 2024;48(5):694–701.

15. Xiang L, Wu H, Pan A, et al. FTO genotype and weight loss in diet and lifestyle interventions: a systematic review and meta-analysis. Am J Clin Nutr. 2016;103(4):1162–1170.

16. Livingstone KM, Celis-Morales C, Papandonatos GD, et al. FTO genotype and weight loss: systematic review and meta-analysis of 9563 individual participant data from eight randomised controlled trials. BMJ. 2016;354:i4707.

17. Gardner CD, Trepanowski JF, Del Gobbo LC, et al. Effect of Low-Fat vs Low-Carbohydrate Diet on 12-Month Weight Loss in Overweight Adults and the Association With Genotype Pattern or Insulin Secretion: The DIETFITS Randomized Clinical Trial. JAMA. 2018;319(7):667–679.

18. Papandonatos GD, Pan Q, Pajewski NM, et al. Genetic Predisposition to Weight Loss and Regain With Lifestyle Intervention: Analyses From the Diabetes Prevention Program and the Look AHEAD Randomized Controlled Trials. Diabetes. 2015;64(12):4312–4321.

19. Kurki MI, Karjalainen J, Palta P, et al. FinnGen provides genetic insights from a well-phenotyped isolated population. Nature. 2023;613(7944):508-518.

20. Rodosthenous RS, Niemi MEK, Kallio L, et al. Recontacting biobank participants to collect lifestyle, behavioural and cognitive information via online questionnaires: lessons from a pilot study within FinnGen. BMJ Open. 2022;12(10):e064695.

21. Harris PA, Taylor R, Minor BL, et al. The REDCap consortium: Building an international community of software platform partners. J Biomed Inform. 2019;95:103208.

22. Willer CJ, Li Y, Abecasis GR. METAL: fast and efficient meta-analysis of genomewide association scans. Bioinformatics. 2010;26(17):2190–2191.

23. Ge T, Chen CY, Ni Y, Feng YA, Smoller JW. Polygenic prediction via Bayesian regression and continuous shrinkage priors. Nat Commun. 2019;10(1):1776.

24. Chang CC, Chow CC, Tellier LC, Vattikuti S, Purcell SM, Lee JJ. Second-generation PLINK: rising to the challenge of larger and richer datasets. Gigascience. 2015;4:7.

