## Supplementary_Figures for "Effect of a Dietary Intervention on Weight Loss in Adults with High vs Low Genetic Predisposition to Higher BMI: A Randomized Diet Intervention Trial"

**SUPPLEMENTARY MATERIAL**

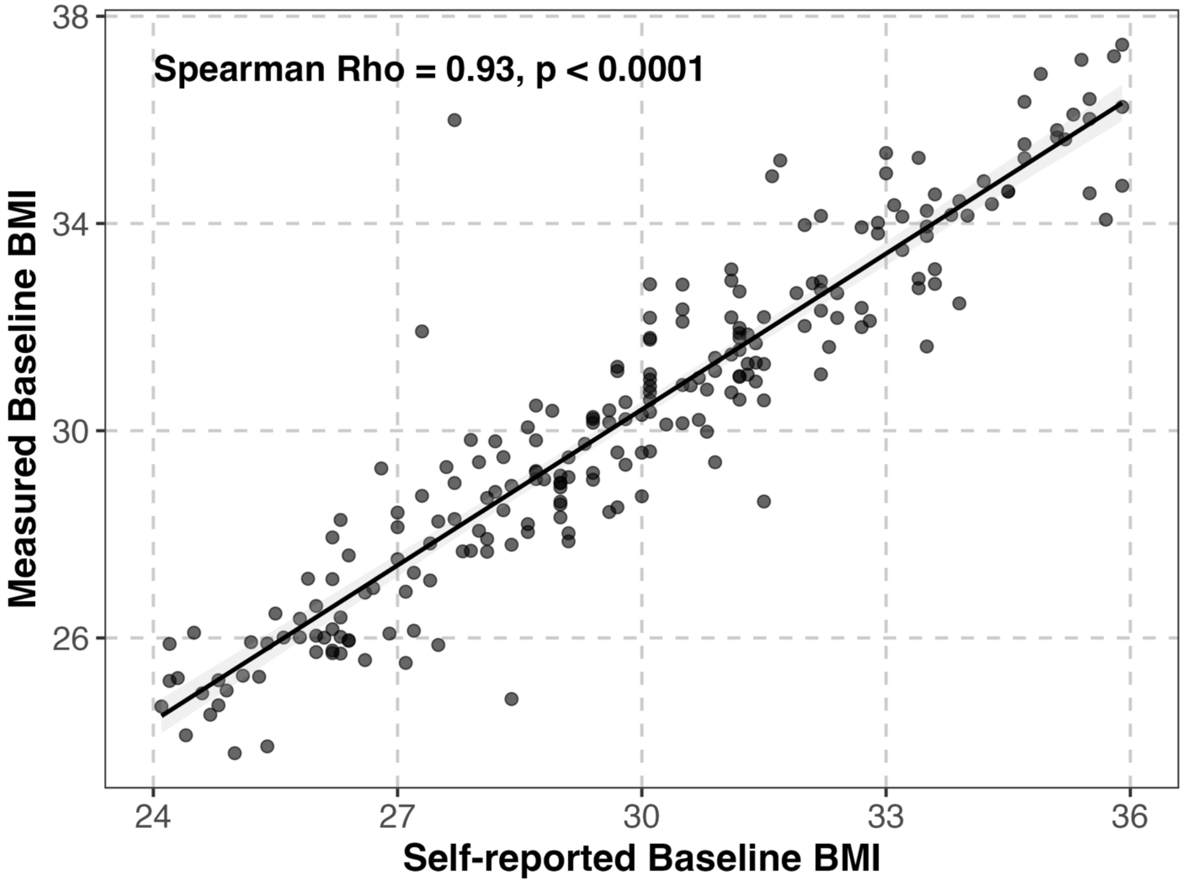


**Supplementary figure 1:** Scatterplot showing the correlation between self-reported and lab-measured BMI.

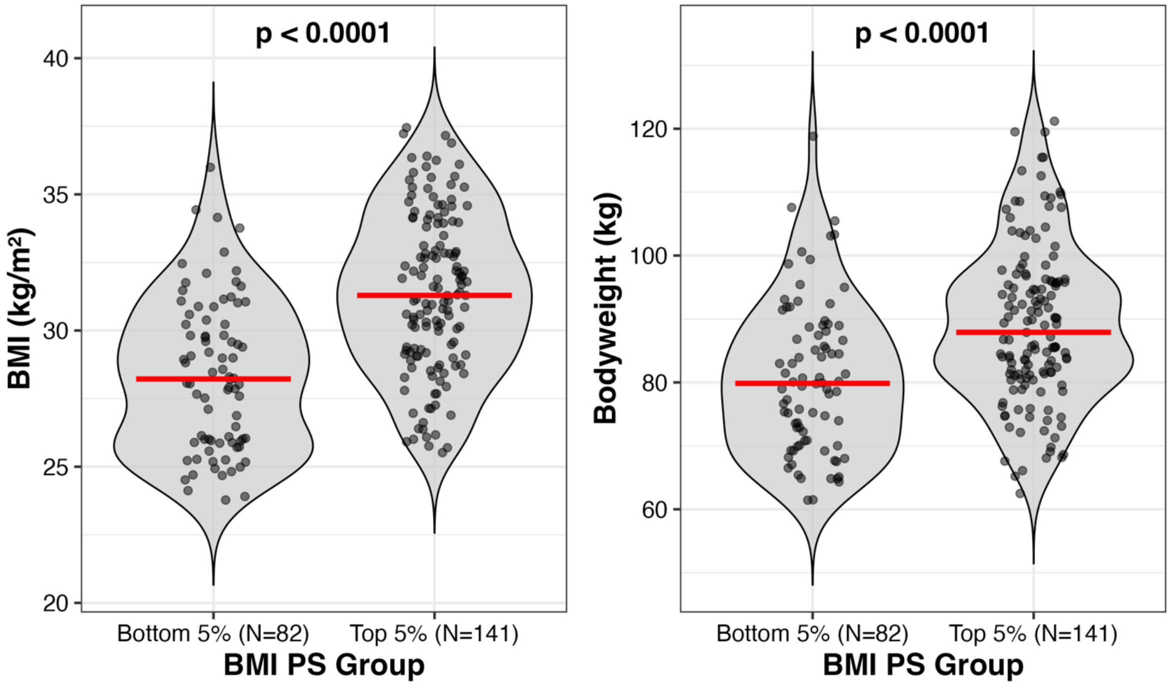


**Supplementary figure 2:** Difference in baseline BMI and body weight among study participants in the Top and Bottom 5% of the BMI PS (N=223). P-values were tested using Mann-Whitney U (Wilcoxon rank-sum) non-parametric test. Red bar shows the median values.

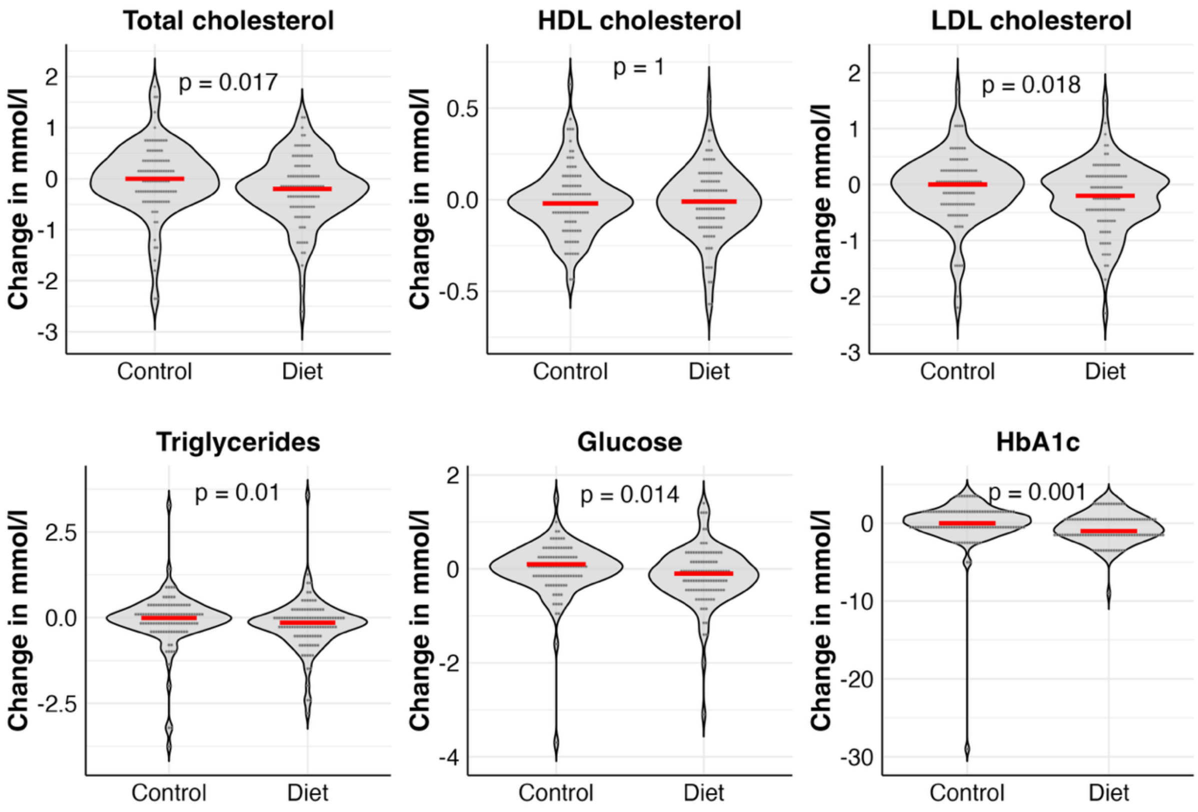


**Supplementary figure 3**: Changes in lab values between the control (N=104) and diet intervention (N=106) groups. P-values were tested using Mann-Whitney U (Wilcoxon rank-sum) non-parametric test. Red bar shows the median values.
