## Supplementary_Material_File_1 for "Effect of a Dietary Intervention on Weight Loss in Adults with High vs Low Genetic Predisposition to Higher BMI: A Randomized Diet Intervention Trial"

### GENEROOS-The role of genetics in weight loss research

#### 1. Background

The prevalence of both overweight and obesity is increasing at an alarming rate worldwide. In fact, it is estimated that 2.16 billion adults (38%) will be overweight and 1.12 billion adults (20%) will be obese by 2030. Among other risk factors (e.g., environmental, behavioral, and medical), genetics holds an important role. The heritability of body mass index (BMI) is moderate to high, with estimates from twin studies ranging from 0.47 to 0.90 [PMID: 22645519]. Genome-wide association studies (GWAS) have discovered hundreds of loci associated with BMI [PMID: 30124842, 25673413]. Polygenic scores (PS) generated from an aggregated combination of millions of genetic variants explain ~6.0% of the variance of BMI. The correlation between PS with actual BMI is ~0.22 [PMID: 30124842]. In the UK biobank, severe obesity - defined as BMI > 40 kg/$m^2$ - was present in 1,621 of 28,784 (5.6%) of those in the top decile of the PS versus 69 of 28,834 (0.2%) of those in the bottom decile, corresponding to a 25-fold gradient in risk of severe obesity ($p < 0.0001$) [PMID: 31002795]. Therefore, PS for BMI is strongly predictive of BMI and obesity risk.

There have been numerous diet intervention studies aimed at reducing body weight in overweight and obese individuals. A meta-analysis of clinical trials showed a mean weight loss of 5 to 8.5 kg (5% to 9%) during the first 6 months from interventions involving a reduced-energy diet with weight plateaus at approximately 6 months [17904936]. Smaller benefits were observed on weight loss maintenance beyond 12 months [25134100] and reduction in all-cause mortality [PMID: 29138133].

Little is known about the effect of genetic background in impacting the effectiveness of diet interventions. Candidate-gene retrospective studies have shown that individuals carrying the homozygous FTO obesity-predisposing allele - the strongest genetic signal for BMI - may lose more weight through diet/lifestyle interventions than noncarriers [PMID: 6888713]. Studies that examined the role of PS in modifying diet intervention effects are scarce. A retrospective analysis of the POUNDS LOST trial suggests that individuals with a lower genetic risk of diabetes may benefit more from consuming a low-protein weight-loss diet in improving insulin resistance [PMID: 27281308]. To our knowledge, the DIETFITS trial is the only prospective trial that has been designed to evaluate the effect of genotypes in modifying the impact of a healthy low-fat (HLF) diet vs a healthy low-carbohydrate (HLC) diet on weight change [PMID: 29466592]. The study, however, does not consider PS.

In this study, we will leverage the unique opportunity provided by the Finnish biobank research to re-contact 1200 individuals who have extreme genetic predisposition for high/low BMI as measured by a PS for BMI and evaluate how the randomized diet intervention effect varies between the two extreme groups.

#### 2. Aim

The aim of the study is to determine whether the BMI polygenic score impacts the effectiveness of dietary/lifestyle intervention in reducing BMI among individuals with elevated BMI. The overview of the proposed study is shown in **Figure 1**. In brief, we will invite overweight (25-35 BMI) individuals with very high (Top 5%; n=600) and very low (Bottom 5%; n=600) PS for BMI who have given biobank consent, have genetic data available through their biobank, and fulfill the study's inclusion criteria (see Section 4). Half of the participants in each group will be randomized to enroll in a dietary/lifestyle coaching intervention lasting 6 months, or will not receive any dietary advice or information. All participants will be asked for two lab visits, one at baseline and one at the end of the study, to have their fasting blood samples collected.

**Figure 1.** An overview of the GENEROOS study.

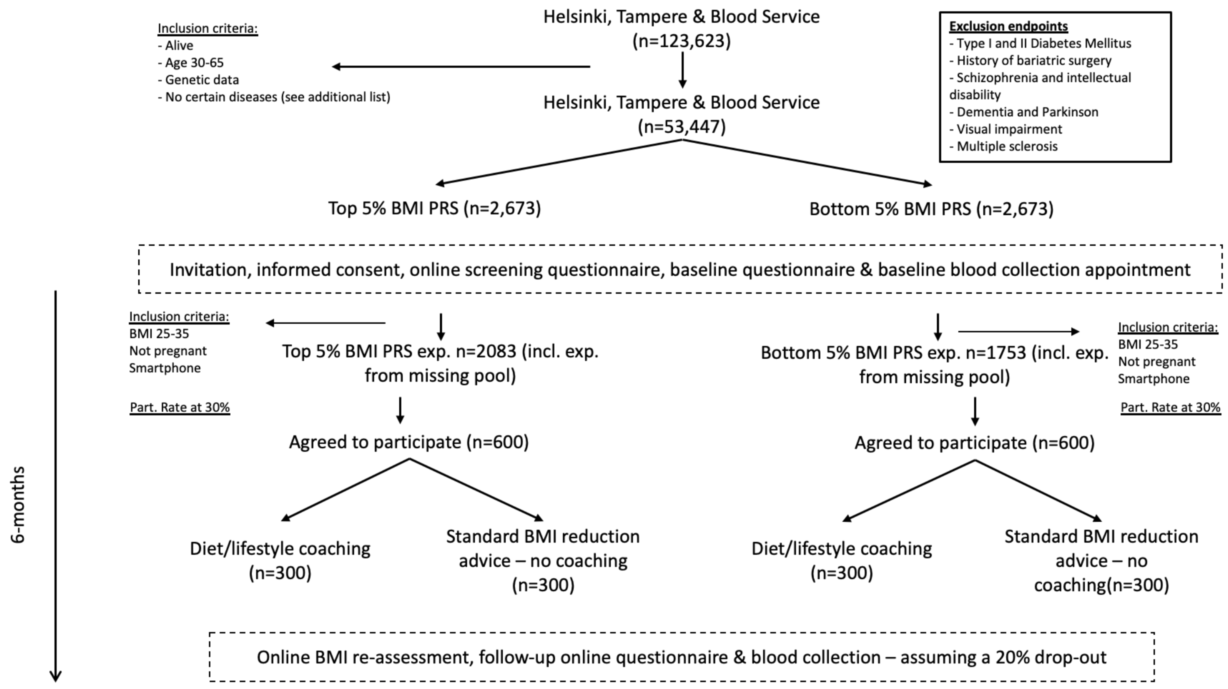


#### 3. Impact

The role of genetic information in identifying individuals who would mostly benefit from diet interventions is not known. To the best of our knowledge, prospective studies assessing this hypothesis are lacking. This well-powered prospective study leverages the unique opportunity to re-contact individuals at the extremes of BMI PS from a large pool of individuals with already available genetic information. This study will determine whether BMI PS can be used to identify overweight and obese individuals who are more likely to succeed in a reduced-energy dietary intervention, which will assist our efforts in curbing the overweight and obesity epidemic.

#### 4. Description of the Study Participants

In this prospective study, we will select individuals from Helsinki, Tampere and the Blood Donor Biobanks who are already enrolled by the larger FinnGen study. Firstly, we will identify eligible individuals based on the following **inclusion criteria**:

1. Age of 30-65 years old
2. BMI of 25-35
3. Bottom 5$^{th}$ percentile and top 95$^{th}$ percentile of the PS for BMI calculated based on the results from Yengo et al [PMID: 30124842] and meta-analyzed with the results from a GWAS of BMI in FinnGen for the participants not included in the studies.
4. Provided a recontacting informed consent to their Biobank. Once the eligible study participants are identified, we will invite them to participate in our study through the participating Biobanks.

We will also consider the following **exclusion criteria**:

1. Diagnosis of both Type I and II Diabetes Mellitus (ICD-10: E10-E14)
2. History of bariatric surgery (Operation codes: JDF00–01, JDF10–11, JDF20–21, JDF 96–97, JFD 03–04, JFD20 and JFD96)
3. Diagnosis of schizophrenia and intellectual disability (ICD-10: F20-F29, F70-F79)
4. Diagnosis of dementia and parkinson (ICD-10: F00-F09, G20, G30)
5. Diagnosis of visual impairment (ICD-10: H54)
6. Diagnosis of multiple sclerosis (ICD-10: G35)

#### 5. Work plan and description of research methods

To identify individuals who are eligible, we will start by estimating the total number of existing Helsinki, Tampere, and Blood Donor biobank-consented individuals (n=123,623). We will then select those individuals who are alive, have genetic data available, are between 30-65 years old, and have not been diagnosed with any of the diseases specified in the exclusion criteria above (n=53,447). Out of those, we will select individuals in the lowest 5$^{th}$ (n=2,673) and highest 95$^{th}$ (n=2,673) percentiles of the PS for BMI. Currently, ~30% of the BMI information is missing from both the highest (773/2763) and lowest (855/2673) PSs. Therefore, after the PS selection, we are planning to send everyone an invitation to participate in our study (except for those we already know their BMI is outside the 23-36 range) and ask them to report their weight and height among other study eligibility questions and given they have provided their electronic consent first (see complete questionnaire in Appendix 1). Finally, we will select individuals with a confirmed BMI between 25-35 (based on their measured and/or self-reported BMI). According to our estimates, we expect to have 2,083 eligible individuals in the highest 95$^{th}$ percentile and 1,753 in the lowest 5$^{th}$ percentile of the PS for BMI in this BMI range. Out of those, we assume (based on our previous experience with the FinnGen recall pilot study) that ~30% will agree to participate in the study, which brings the numbers to ~600 participants for each arm (total of 1,200) of the study. For each PS arm, we will randomize participants into two equal groups based on the type of dietary intervention they will receive **(Figure 1)**.

*5.1. Re-contacting individuals via their biobanks*

Eligible individuals will be re-contacted by their Biobank, namely Helsinki, Tampere Biobank, and Blood Service. More specifically, we will provide a list of FinnGen IDs for all eligible individuals to the participating Biobanks, who will then convert it to social security identifiers to confirm that selected individuals are alive, have not relocated, and have a valid biobank consent for re-contacting. Once the list is validated, the Biobanks will additionally apply any ICD-10 disease exclusions (see Section 4), and then invite the eligible individuals to participate in our study. Invitations will be sent to all eligible individuals via a physical letter, unless an individual had specified otherwise, and will include the invitation letter and bulletin of participants, where they can find additional information about the study. Individuals who are interested in participating will be asked to log in online in the study portal (https://omabiopankki.fingenious.fi/login), provide an electronic informed consent, and complete the eligibility survey, which includes reporting of their height and body weight for BMI calculation and other questions **(see Appendix 1)**. If the calculated BMI is within the 25-35 range, and they have passed the other eligibility criteria, they will be allowed to enroll the study, complete a short online questionnaire (see section 5.2), and book an appointment for their visit to a local clinical laboratory (e.g. HUSLAB in Helsinki) for the collection of their baseline fasting blood sample within 4 weeks from their enrollment. To validate the self-reported BMI measurements, we will coordinate with one site (i.e., Tampere Biobank) and have a research nurse measure participants' BMI when they come into the lab for blood collection. This will be done for both visits, and BMI measurements will be correlated with the provided self-reported values.

*5.2. Online questionnaires*

Participants who meet all the criteria and are eligible to participate in the study will be asked to complete a baseline online questionnaire. The questionnaire is built in RedCap and will be accessible via a link in each participant's personal OmaBiopankki page. The questionnaire is designed to be short (max 15min to complete) and cover basic demographic questions (e.g., age, sex, marital status, etc), as well as questions related to eating habits and psychological eating. At the end of the study, we will ask the participants to complete the same questionnaire as a follow-up. The complete online questionnaire is shown in **Appendix 2**.

*5.3. Blood sample collection and biomarker analysis*

Whole blood samples (2 x 10 mL) will be collected by a trained nurse in EDTA tubes at each participating lab. All blood samples will be shipped to a central lab (same day delivery) where biomarker analyses (e.g., lipids, HbA1C, glucose) will be performed. The coordination of the sample collection and analysis will be done by the biobanks. The results of biomarker analysis (i.e., at baseline and follow-up) will be returned to study participants who provided informed consent and agreed to receive them after the end of the study.

*5.4. Measurement of primary outcome: BMI*

We will measure BMI four times during the study; one at baseline, one at the end (month 6), and two more times every two months from enrollment (month 2 and 4). Baseline BMI will be calculated using self-reported body weight and height via the eligibility survey at enrollment.

Following enrollment, BMI will be further validated by having a trained nurse measure participants' body weight and height during their first visit to the closest clinic to have their blood drawn. BMI validation will only be done at the Tampere site, which will serve as our pilot site. We will also reach out every two months to the participants, asking them to report their BMI via an online questionnaire administered via the Omabiopankki portal (see **Appendix 3**). Participants will be advised to weigh themselves in the morning, before breakfast, and after emptying their bladders. In addition, participants will be instructed to measure under the same conditions over time.

*5.5. Diet intervention*

Half of the participants (50%) in each of the two groups (top & bottom 5% of BMI PS) are randomly assigned to the diet coaching or control group. Following blood collection at baseline, participants in the diet coaching group will be contacted by ViaEsca, our external collaborator, and informed about the dietary intervention plan. In addition, participants in the diet coaching group will receive a 120 EUR coupon and be advised to order a wearable tracking device before the initiation of their program. ViaEsca will be blinded as to whether the study participants belong to the high or low PS group. Detailed material and support will be provided to the intervention arm of our study, while no information will be provided to our control arm.

The ViaEsca intervention: The main focus is to guide participants to lower their body weight, which will subsequently lower their BMI as well. An equally important aim of the study is to help participants permanently keep a balanced way of eating; this balance is based on a combination of calorie intake/burning and balanced nutrition based on the official nutritional guidelines. To achieve that, all participants of this intervention will get the same information, training and support at the beginning of the program, which will ensure that everyone has the same starting point.

- Food plan: Everybody gets 5 meals/day based on their calorie needs minus 500 kcal/day less than their daily calorie expenditure. All meals will be tracked via the ViaEsca mobile application, including any meals chosen by the participants that are outside the recommended food plan and the calories will be calculated automatically based on the information provided.
- Activity: To calculate the calories burned from any activities, we will give all participants of this arm (n=600) an activity tracking device. The first calorie level is calculated based on the information in the registration form: age, height, weight, gender, personal view of the daily activity. After that, and once the activity tracker is synced to the program, the calorie level is calculated based on the 7 days average calories burned.
- Communication: All participants will have an initial onboarding call with one of ViaEsca's coaches. A follow up call will also be had 3 weeks after the start of their program. In addition, participants will be encouraged to contact their coach for any questions or concerns they may have via a text, which is a built-in function in ViaEsca's phone application.

*5.6. Follow-up visit and wearable device for the control group*

The duration of this diet intervention study is six months. In addition to the baseline online questionnaire and blood sampling, we will repeat this process at the end of the study. All participants will be asked to answer the same online questionnaire and visit the same lab for their second fasting blood sample collection. Once they complete both the online questionnaire and blood sample collection, we will provide the opportunity to all participants to learn about which PS group they are (low or high PS for BMI) by sending us an email or giving us a call. Last, we will provide the same 120 EUR coupon for the wearable device to all participants in the control group.

The overall timeline of the study is shown in Figure 2 below.

**Figure 2.** The overall timeline of the GENEROOS study.

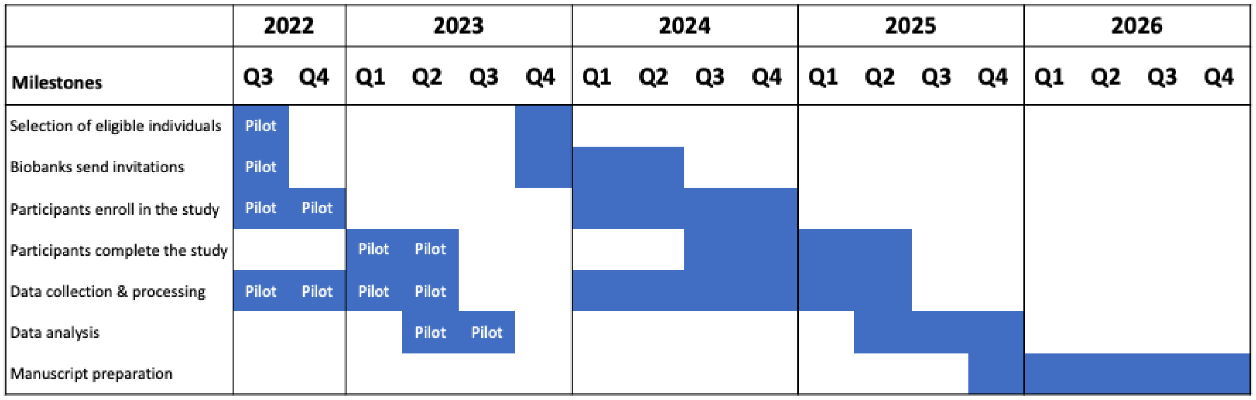


#### 6. Data processing and data management

All data collected in this study will be transferred and stored at CSC's ePOUTA secure server, which is developed for processing sensitive data. For the processing and analysis of data, CSC's secure computing environment will be used by authorized researchers who are granted data processing access. Before the end of the study (and erasure of all the data), any data that may need to be transferred to the participating Biobanks will be coordinated by the research team. The project will end on 31/12/2026, at which time all personal-level data will be erased. By this date, we are expecting to have gathered, analyzed, and published the findings from this study in a relevant peer-reviewed journal (e.g., BMJ, JAMA, NEJM, or other). A detailed description of data processing and data management processes can be found in the Data Protection Impact Assessment document.

#### 7. Data analysis plan and power calculations

The main hypothesis that we will test in the study is that the effect of the diet intervention differs between the two groups (the lowest 5$^{th}$ vs the highest 95$^{th}$). That is, we are testing the interaction between the group indicator and the diet indicator in their association with BMI differences between baseline and after 6 months (ΔBMI). We will test this interaction using a linear model such as: ΔBMI ~ diet_group*PRS_tail + age + age^2 + sex + center.

The secondary hypothesis is that the diet intervention reduces BMI. To test this hypothesis, we will perform a linear model such as: ΔBMI ~ diet_group + age + age^2 + sex + center.

*Power calculations*

We applied the selection criteria described above to FinnGen R7. The BMI difference between the two PRS tails was 1.9 (the mean BMI was 27.8 for the lowest $5^{th}$ and 29.7 for the highest $95^{th}$). 55% are women and 45% are men. We use this information to perform power calculations.

We assume that the diet intervention has a baseline effect of 5%, which is consistent with what is reported in the literature and considered to be clinically significant [PMID: 9347414, 19127177]. The power calculation (**Figure 3**) tests how being in the two groups based on PS modifies the expected 5% diet effect. We test this effect to be symmetrical. For example, a ± 2% should be interpreted as a 7% diet effect in the highest $95^{th}$ percentile and a 3% diet effect in the lowest $5^{th}$. We have 80% power to detect a ± 2% or higher difference in diet reduction between the two groups based on PS. For example, if we consider an individual with height=1.70 m and weight=87 kg (BMI=30.1), we have the power to detect a 6.1 kg loss in the highest $95^{th}$ vs a 2.5 kg loss in the lowest $5^{th}$.

**Figure 3**: Power calculations.

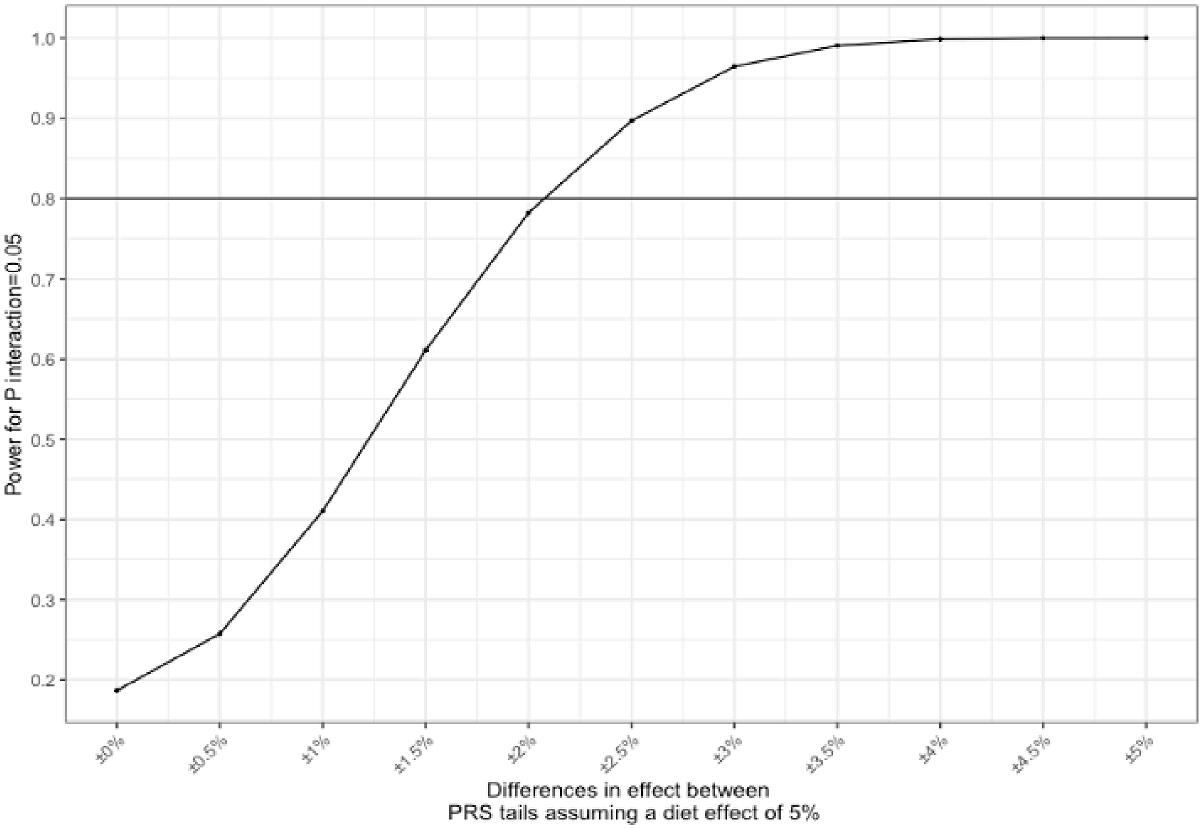


**8. Research team**

The core research team consists of Drs. Andrea Ganna (Principal Investigator), Kirsi Pietiläinen (Collaborator), Elisabeth Widen (Collaborator), Samuli Ripatti (Collaborator), Rodosthenis Rodosthenous (Research Coordinator), and Anne Carson (Project Coordinator). In addition to the research team, the study will be supported by one project coordinator and two experienced research nurses at each of the recruitment sites. Last, the study will be supported by a FINBB project manager who will oversee coordination between the involved biobanks and the research team, as well as an IT support who will ensure the functionality of the OmaBiopankki portal during the study.

**9. Funding**

The GENEROOS study is fully funded by the European Research Council (ERC-2020-STG), AI-PREVENT 945733 (PI Andrea Ganna). The proposal is titled “A nationwide artificial intelligence risk assessment for primary prevention of cardiometabolic diseases.”
