## Supplementary_Material_File_3 for "Effect of a Dietary Intervention on Weight Loss in Adults with High vs Low Genetic Predisposition to Higher BMI: A Randomized Diet Intervention Trial"

**Pre-screening questionnaire (online in RedCap)**

All invited study subjects will be asked to fill in the information requested below to confirm their eligibility to participate in the GENEROOS study.

1. Date of birth (dd-mm-yyyy) ______________

2. Body weight (kg) _____________

3. Height (cm) ____________

*Based on each subject's body weight and height values, their BMI will be automatically calculated and shown as follows:*

4. Your body mass index (BMI) is: ___________

5. Are you pregnant, or have you delivered a baby in the past 6 months?
   ( ) Yes
   ( ) No

6. Do you have a smartphone?
   ( ) Yes
   ( ) No

7. Are you interested in losing weight?
   ( ) Not interested
   ( ) Somewhat interested
   ( ) Very interested

At the end of the questionnaire, each person will receive a message about whether they are eligible to participate in the study or not, according to the following inclusion criteria:
- Age between 30-65 years
- BMI between 23-36
- The person is NOT pregnant or has delivered a baby in the last 6 months
- The person owns a smartphone
- The person is not very interested or somewhat interested in losing weight

If the person who answered the questionnaire is eligible, they will be notified with the message, "Congratulations, you are eligible to join the GENEROOS study. Please proceed to the e-consent page."

If the person is not eligible, they will be notified with the following text:
"Unfortunately, you are not eligible to join the GENEROOS study because, according to your responses, you:
(1) are outside the 30-65 years of age range, **OR**
(2) have a BMI below 23 or over 36, which is outside the focus of this study, **OR**

(3) are pregnant or have delivered a baby in the past six months, **OR**
(4) do not have a smartphone, **OR**
(5) are not interested in losing weight

Thank you for your interest and time in completing this questionnaire!”
